# Red blood cell distribution as Potential Predictor of Mortality in Diabetic Foot Patients

**DOI:** 10.1101/2024.09.25.24314391

**Authors:** Chunmei Gou, Siyi Huang, Li Huang, Tinggang Wang, Guangtao Huang

## Abstract

**Background:** Red blood cell distribution width (RDW) reflects the heterogeneity of red blood cell volume, which reflects the variable width of red blood cell (RBC). RDW has been proved as predictor of mortality among several diseases. The purpose of this study is to analyze the relationship between RDW and mortality of diabetic foot patients.

**Methods:** We first collect clinic data from the public database MIMIC-III. Kruskal Wallis rank sum test was used to analyze the association between RDW and DF mortality, and to evaluate the relationship between them. Univariate and multivariate logistic regression analysis was used for determining the risk factors and prognosis of DF patients.

**Results:** A total of 283 patients were included in this study, with an average age of 64.0 [54.0,70.5] years, including 193 males and 90 females. We divided RDW into three groups (high, moderate and low) according to RDW tertiles and then compared the mortality of the three groups. The high RDW group (RDW > 16.8%) had significant higher mortality (P = 0.031). In multivariate logistic regression analysis, RDW, SOFA score and APS are risk factors for death in diabetic foot. After adjusting for confounding factors in model II, RDW remains a particularly strong predictor of mortality.

**Conclusions:** A total of 283 patients were included in this study, with an average age of 64.0 [54.0,70.5] years, including 1We confirm that RDW is an independent predictor of mortality in DF patients,and the higher the RDW, the higher the mortality of DF patients.

## Introduction

In recent years, the prevalence of diabetes is rising. Among them, diabetic foot is one of the most important complications^1^. Diabetic foot constitutes a major health and economic burden, which is one of the main reasons for hospitalization among diabetic patients^2-5^. Up to 25% of diabetic patients might suffer from foot ulcers during lifelong time. The annual incidence rate of diabetic foot is 2.4 - 2.6% and the prevalence rate is around 4 - 10%^6-10^. The early symptoms of diabetic foot are usually numbness, which are easy to be ignored by patients and clinicians. Meanwhile, DF is accompanied by many complications, leading to a serious decline in the quality of life of patients, which can lead to amputation or even death^8, 9, 11-16^. Diabetic patients with DF ulcers have twice as much mortality as those without ulcers^17^.

At present, the commonly used clinical treatment strategies include blood glucose control, dressing, debridement, negative pressure wound therapy and so on^18-23^. However, there are still many diabetic foot patients come out with poor prognosis. Considering the high incidence and mortality of diabetic foot, simple and feasible indicators are urgently needed to forecast the mortality of diabetic foot and ameliorate the prognosis of patients.

Some studies have evaluated the current predictors of diabetic foot mortality. In multivariate analysis, peripheral artery disease, procalcitonin, microbial growth in deep tissue and renal impairment are independent risk factors for death^24-26^. RDW refers to the variation degree of red blood cell volume. The larger the value, the greater the morphological divergence of red blood cells in blood sample. It is usually seen in various anemia, abnormalities of hematopoietic system or congenital red blood cell abnormalities^27^. Recently, RDW has been associated with mortality of several diseases^28^, such as heart failure, brain infarction, COVID-19, acute kidney injury, severe acute pancreatitis (SAP), stroke and community-acquired pneumonia^29-35^. Up to now, there is no relevant research on RDW and DF mortality. In this study, RDW is explored for predicting the mortality of DF, which is of great importance for prognosis of patients.

## Methods

### The Database

Our study is based on the Multiparameter Intelligent Monitoring in Intensive Care Database III version 1.3 (MIMIC-III V1.3). It is a free public resource. It includes more than 40000 ICU patients from 2001 to 2012, all of whom were admitted to Beth Israel Deaconess Medical Center (Boston, Massachusetts, USA). The database was established by the Massachusetts Institute of Technology (MIT, Cambridge, MA, USA) and Beth Israel Deaconess Medical Center. Since MIMIC-3 is publicly available and deidentified, the secondary analysis in this study is exempt from the institutional review boards (IRBs) of Beth Israel Deaconess Medical Center and the Massachusetts Institute of Technology review. As a result, this study did not undergo an IRB review. According to the patient privacy protection act, the identity of all patients is unknown.

### Population Selection Criteria

In total, 283 people were included. The diabetic foot patients were older than 18 years old and hospitalized for more than two days were included. If the patient has one of the following conditions, he will be excluded: (1) RDW value was not measured during hospitalization; (2) patients with hematological diseases; (3) personal data loss > 5%.

### Data Extraction

The data we obtained are from MIMIC-III. The extracted data include physiological information such as heartrate, respiratory rate, o2saturation, nibp_systolic, nibp_diastolic and temperature acquired by bedside monitor. Age, gender and laboratory parameters were also included. We extracted the following laboratory parameters: body mass index (BMI); creatinine; alanine aminotransferase (ALT); aspartate aminotransferase (AST); bicarbonate; hematocrit (HCT); urea nitrogen (BUN); serum calcium; serum sodium; serum potassium; white blood cell count (WBC); neutrophils; hemoglobin; platelet count (PLT); alkaline phosphatase; bilirubin; albumin and glucose. Osteomyelitis, amputation, SOFA and APS were also calculated to valuate the prognosis of the disease. The outcome measure was hospital mortality. Baseline features were extracted 24 hours after admission.

### Regression analysis and subgroup analysis

Univariate and multivariate logistic regression analysis were used for determining the risk factors of clinical outcomes. Through multiple regression, we designed two models to determine the factors related to RDW value. In model I, we adjusted covariates for age and gender. In model II, we further adjusted covariates for age, gender, body mass index, osteomyelitis, amputation, bicarbonate, hematocrit, oxygen saturation, white blood cell count, neutrophils, alkaline phosphatase, bilirubin, albumin and temperature. We would use the above data to explore the relationship between RDW and diabetic foot mortality, and use corrected odds ratio (OR) and 95% confidence interval. We also performed subgroup analysis to comfirm whether RDW has different effects on different subgroups classified by age, gender, HCT, HGB, SOFA, APS, ect.

### Statistical Analysis

All patients obtained were divided into three groups according to RDW tertiles. When p < 0.05, the difference was statistically significant. Continuous variables were represented by mean ± standard deviation (SD) and contrasted by ANOVA or Kruskal Wallis test.

## Results

### Subject Characteristics

We divided RDW into three groups (Table 1). In the low-RDW group (11.5 < RDW< 14.5), there were 90 (31.8%) patients. There were 98 (34.6%) and 95 (33.6%) patients in moderate-RDW group (14.6 <RDW < 16.5) and high-RDW group (16.6 < RDW < 24), respectively. The grouping and laboratory data of RDW are shown in table 1.

**Table 1:** Population distribution characteristics of patients with RDW.

| N=283 | RDW |  |  | P |
| --- | --- | --- | --- | --- |
|  | 11.5-14.5 | 14.6-16.5 | 16.6-24 |  |
|  | 90 (31.8%) | 98 (34.6%) | 95 (33.6%) |  |
| rdw | 13.7 (13.0-14.0) | 15.6 (15.0-16.0) | 18.4 (17.3-20.1) | <0.001 |
| age | 63.0 (52.2-69.0) | 64.0 (54.5-73.8) | 63.0 (54.0-68.5) | 0.174 |
| gender |  |  |  | 0.013 |
| male | 68 (75.6%) | 73 (74.5%) | 55 (57.9%) |  |
| female | 22 (24.4%) | 25 (25.5%) | 40 (42.1%) |  |
| osteomyelitis | 11 (12.2%) | 9 (9.2%) | 11 (11.6%) | 0.778 |
| amputation | 4 (4.4%) | 17 (17.3%) | 14 (14.7%) | 0.019 |
| bmi | 29.8 (25.6-36.4) | 30.6 (25.4-37.2) | 28.7 (24.3-34.1) | 0.337 |
| icu_los_hours | 43.0 (23.0-76.0) | 57.0 (34.2-104.8) | 64.0 (25.5-102.0) | 0.893 |
| heartrate | 89.9 (78.0-102.0) | 87.5 (77.2-97.0) | 89.9 (80.0-101.0) | 0.541 |
| Respiratory rate | 19.7 (16.0-22.0) | 19.7 (16.0-23.0) | 19.0 (15.5-22.0) | 0.936 |
| o2saturation | 96.8 (96.8-99.0) | 97.0 (96.8-98.8) | 98.0 (96.0-100.0) | 0.411 |
| nibp_systolic | 122.0 (109.2-140.8) | 121.8 (107.2-138.0) | 112.0 (100.5-128.5) | 0.044 |
| nibp_diastolic | 64.7 (61.0-75.8) | 62.0 (53.2-67.8) | 63.0 (54.0-70.5) | 0.013 |
| temperature | 36.9 (36.5-37.4) | 36.8 (36.5-37.2) | 36.6 (36.3-37.0) | 0.746 |
| creatinine | 1.2 (1.0-1.8) | 1.9 (1.1-3.2) | 2.7 (1.1-4.9) | <0.001 |
| bun | 26.0 (17.5-42.2) | 37.6 (24.0-51.0) | 34.0 (20.5-48.0) | 0.022 |
| alt | 26.5 (15.0-62.3) | 39.0 (17.0-62.3) | 30.0 (15.5-62.3) | 0.054 |
| ast | 23.5 (15.0-90.8) | 58.5 (20.2-90.8) | 35.0 (20.0-90.8) | 0.01 |
| bicarbonate | 24.0 (20.0-27.0) | 23.2 (19.4-26.0) | 23.2 (21.0-26.0) | 0.886 |
| hct | 34.8 (32.1-38.3) | 31.3 (27.5-36.0) | 29.9 (25.1-34.0) | <0.001 |
| calcium | 8.8 (8.4-9.4) | 8.6 (8.1-8.9) | 8.5 (8.0-8.9) | 0.001 |
| sodium | 134.0 (131.0-137.0) | 134.0 (131.0-137.0) | 137.0 (133.5-139.5) | <0.001 |
| potassium | 4.2 (3.9-4.6) | 4.4 (3.8-5.0) | 4.3 (3.9-4.6) | 0.053 |
| wbc | 13.0 (9.8-17.5) | 15.4 (11.4-19.4) | 11.3 (8.4-16.6) | 0.439 |
| neutrophils | 78.8 (74.0-85.0) | 78.8 (78.8-85.8) | 78.8 (76.2-81.5) | 0.396 |
| hgb | 11.6 (10.6-12.9) | 10.4 (8.9-11.8) | 9.6 (7.8-10.8) | <0.001 |
| platelets | 276.0 (192.2-366.2) | 280.6 (234.2-342.5) | 240.0 (172.5-317.0) | 0.105 |
| alkaline_phos | 121.0 (90.5-178.3) | 158.0 (100.5-178.3) | 178.3 (105.0-178.7) | 0.01 |
| bilirubin | 0.7 (0.4-1.1) | 0.8 (0.5-1.1) | 1.1 (0.5-1.1) | 0.055 |
| albumin | 2.6 (2.5-3.1) | 2.6 (2.3-2.7) | 2.6 (2.4-2.8) | 0.004 |
| glucose | 207.5 (136.2-369.8) | 201.5 (147.0-263.0) | 128.0 (105.0-199.5) | <0.001 |
| sofa | 4.0 (1.0-5.0) | 5.0 (3.0-7.0) | 7.0 (5.0-9.0) | <0.001 |
| aps | 52.0 (41.0-59.5) | 59.5 (53.0-73.2) | 59.5 (45.0-75.0) | 0.013 |

There were no significant differences in age and BMI among the groups. At the same time, we found that the group with higher RDW had lower blood pressure. Patients with higher RDW had a higher risk of amputation. Higher RDW also had higher BUN, creatinine, alkaline , sodium than those with lower RDW. Instead, in the group with high RDW, hematocrit, hemoglobin, calcium and glucose were lower than those in other groups. The high RDW group had a higher SOFA and APS than the low RDW group, indicating that patients with a high RDW were more severely get ill.

### RDW Levels and Mortality

We further divided the patients into survival group and non-survival group. We found that the RDW value of death group was higher than of survival group (15.4% vs 16.8%, P = 0.031). The values of SOFA and APS in the death group were also higher than those in the survival group (Table 2). Furthermore, in univariate logistic regression analysis, many variables are associated with mortality, including RDW, SOFA score and APS (Table 3). The association between RDW and mortality are then confirmed by two multivariate models. Among in model I, RDW existed as a especially strong predictor in patients with diabetic foot (95% ORs: 1.0 - 1.4, P<0.05). After adjustment for age, gender, BMI, osteomyelitis, amputation, bicarbonate, HCT, oxygen saturation, WBC, neutrophils, alkaline phosphatase, bilirubin, albumin, PLT and temperature (model II), RDW still had a strong prediction effect (95% ORs: 1.0 - 1.5, P< 0.05) (Table 4).

**Table 2:**
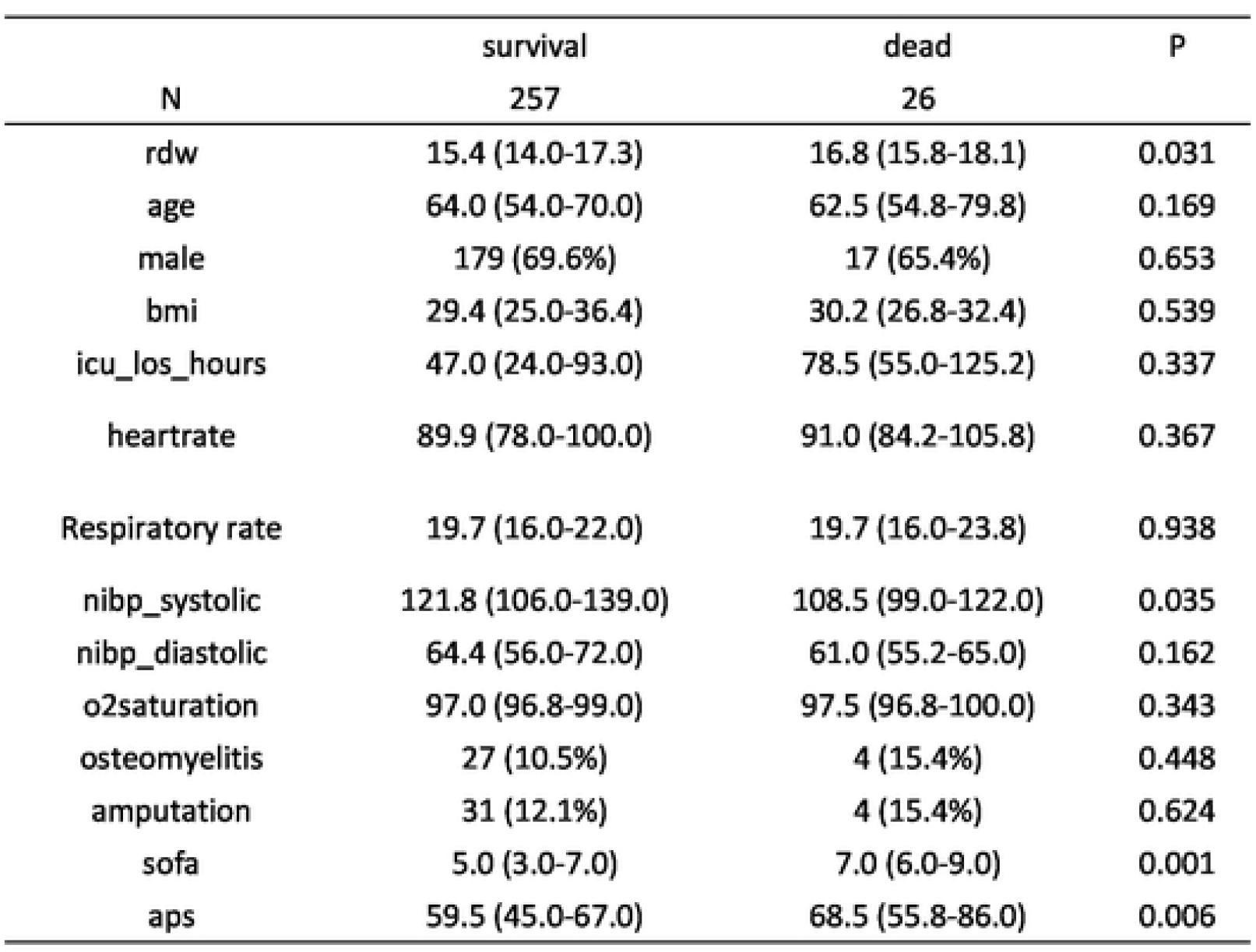
comparison of patient characteristics between survival group and death group.

**Table 3:** univariate factor analysis related to in-hospital death.

|  | Statistics | ORs(95%) | P |
| --- | --- | --- | --- |
| rdw | 15.7 (14.1-17.3) | 1.2 (1.0, 1.4) | **0.034** |
| age | 64.0 (54.0-70.5) | 1.0 (1.0, 1.1) | 0.169 |
| bmi | 29.7 (25.1-36.0) | 1.0 (0.9, 1.0) | 0.537 |
| icu_los_hours | 50.0 (25.5-96.0) | 1.0 (1.0, 1.0) | 0.372 |
| heartrate | 89.9 (79.0-101.0) | 1.0 (1.0, 1.0) | 0.366 |
| respiratoryrate | 19.7 (16.0-22.0) | 1.0 (0.9, 1.1) | 0.937 |
| o2saturation | 97.0 (96.8-99.0) | 1.2 (1.0, 1.4) | 0.12 |
| nibp_systolic | 121.8 (105.0-138.0) | 1.0 (1.0, 1.0) | 0.036 |
| nibp_diastolic | 64.4 (56.0-72.0) | 1.0 (1.0, 1.0) | 0.159 |
| temperature | 36.8 (36.4-37.2) | 1.0 (0.8, 1.1) | 0.74 |
| alt | 30.0 (16.0-62.3) | 1.0 (1.0, 1.0) | 0.75 |
| ast | 34.0 (18.5-90.8) | 1.0 (1.0, 1.0) | 0.511 |
| bicarbonate | 23.2 (20.0-26.0) | 0.9 (0.9, 1.0) | 0.11 |
| hct | 32.2 (27.6-37.2) | 1.0 (0.9, 1.0) | 0.422 |
| calcium | 8.6 (8.2-9.1) | 0.7 (0.5, 1.2) | 0.267 |
| sodium | 135.0 (132.0-138.0) | 1.0 (0.9, 1.1) | 0.938 |
| potassium | 4.3 (3.9-4.8) | 1.3 (0.9, 2.1) | 0.176 |
| wbc | 13.4 (9.4-18.0) | 1.0 (0.9, 1.0) | 0.695 |
| neutrophils | 78.8 (76.2-84.7) | 1.0 (0.9, 1.0) | 0.546 |
| hgb | 10.6 (8.9-12.1) | 0.9 (0.7, 1.1) | 0.156 |
| platelets | 261.0 (194.5-339.0) | 1.0 (1.0, 1.0) | 0.517 |
| alkaline_phos | 153.0 (97.0-178.3) | 1.0 (1.0, 1.0) | 0.8 |
| bilirubin | 0.9 (0.5-1.1) | 1.0 (0.7, 1.3) | 0.762 |
| albumin | 2.6 (2.4-3.0) | 0.6 (0.3, 1.3) | 0.217 |
| glucose | 184.0 (120.0-263.0) | 1.0 (1.0, 1.0) | 0.131 |
| sofa | 5.0 (3.0-7.0) | 1.2 (1.1, 1.4) | **0.002** |
| aps | 59.5 (45.5-68.0) | 1.0 (1.0, 1.0) | **0.008** |
| male | 196 (69.3%) | 1.2 (0.5, 2.8) | 0.654 |
| osteomyelitis | 31 (11.0%) | 1.5 (0.5, 4.8) | 0.451 |
| amputation | 35 (12.4%) | 1.3 (0.4, 4.1) | 0.625 |

**Table 4:**
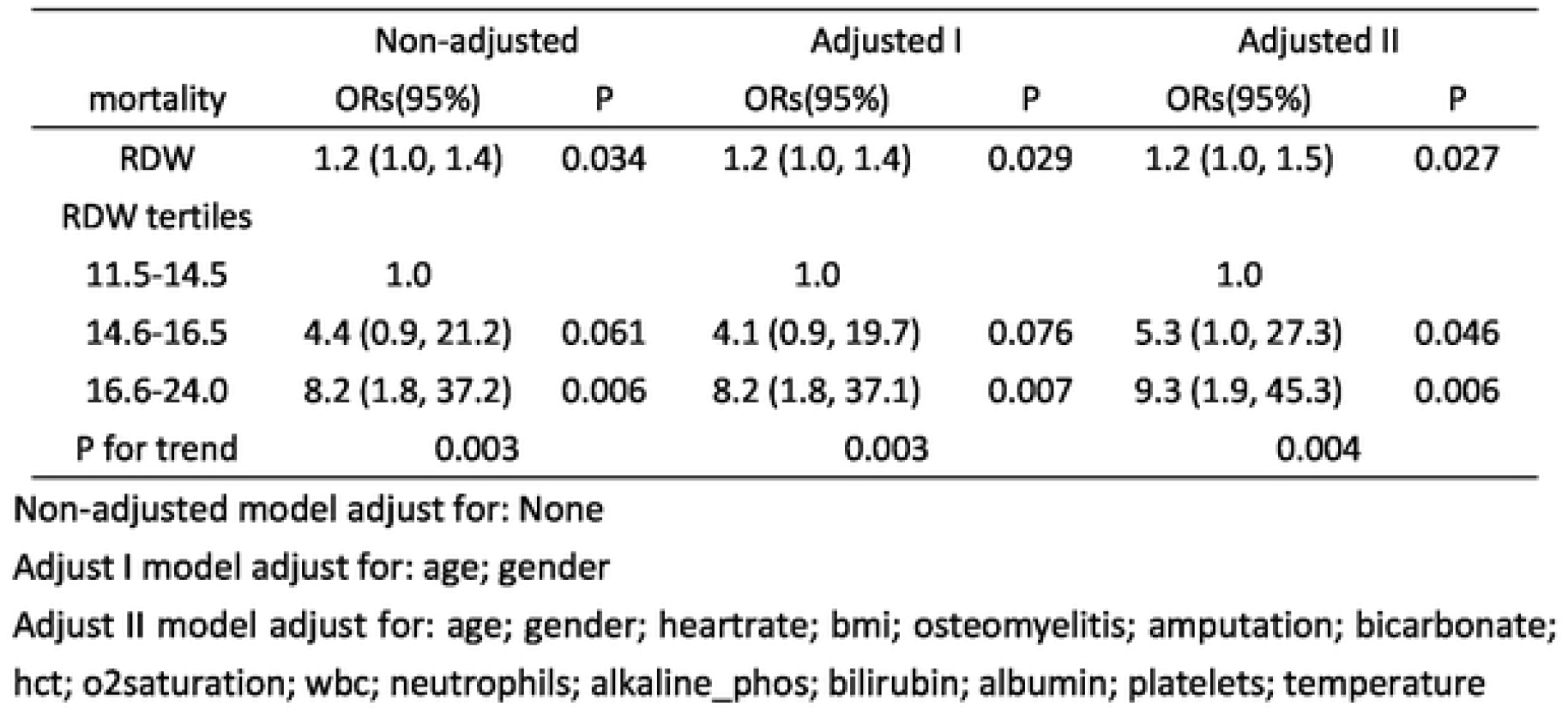
multivariate analysis of in-hospital death and RDW.

| mortality | Non-adjusted |  | Adjusted I |  | Adjusted II |  |
| --- | --- | --- | --- | --- | --- | --- |
|  | ORs(95%) | P | ORs(95%) | P | ORs(95%) | P |
| RDW | 1.2 (1.0, 1.4) | 0.034 | 1.2 (1.0, 1.4) | 0.029 | 1.2 (1.0, 1.5) | 0.027 |
| RDW tertiles |  |  |  |  |  |  |
| 11.5-14.5 | 1.0 |  | 1.0 |  | 1.0 |  |
| 14.6-16.5 | 4.4 (0.9, 21.2) | 0.061 | 4.1 (0.9, 19.7) | 0.076 | 5.3 (1.0, 27.3) | 0.046 |
| 16.6-24.0 | 8.2 (1.8, 37.2) | 0.006 | 8.2 (1.8, 37.1) | 0.007 | 9.3 (1.9, 45.3) | 0.006 |
| P for trend | 0.003 |  | 0.003 |  | 0.004 |  |

Non-adjusted model adjust for: None
Adjust I model adjust for: age; gender
Adjust II model adjust for: age; gender; heartrate; bmi; osteomyelitis; amputation; bicarbonate; hct; o2saturation; wbc; neutrophils; alkaline\_phos; bilirubin; albumin; platelets; temperature

### Subgroup Analysis

We carried out a subgroup analysis to test each indicator that might predict the mortality of diabetic foot (Figure 1). After analysis, we found that HCT, HGB and alkaline are statistically significant in predicting mortality among subgroups. When HCT≥32.2%(OR1.4, 95%CI 1.1 - 1.9), HGB≥10.5g/dl (OR1.6, 95%CI 1.2 - 2.2) and alkaline <153U/L (OR1.4, 95%CI 1.1 - 1.8), RDW was more valuable in predicting the risk of death.

**Figure 1.**
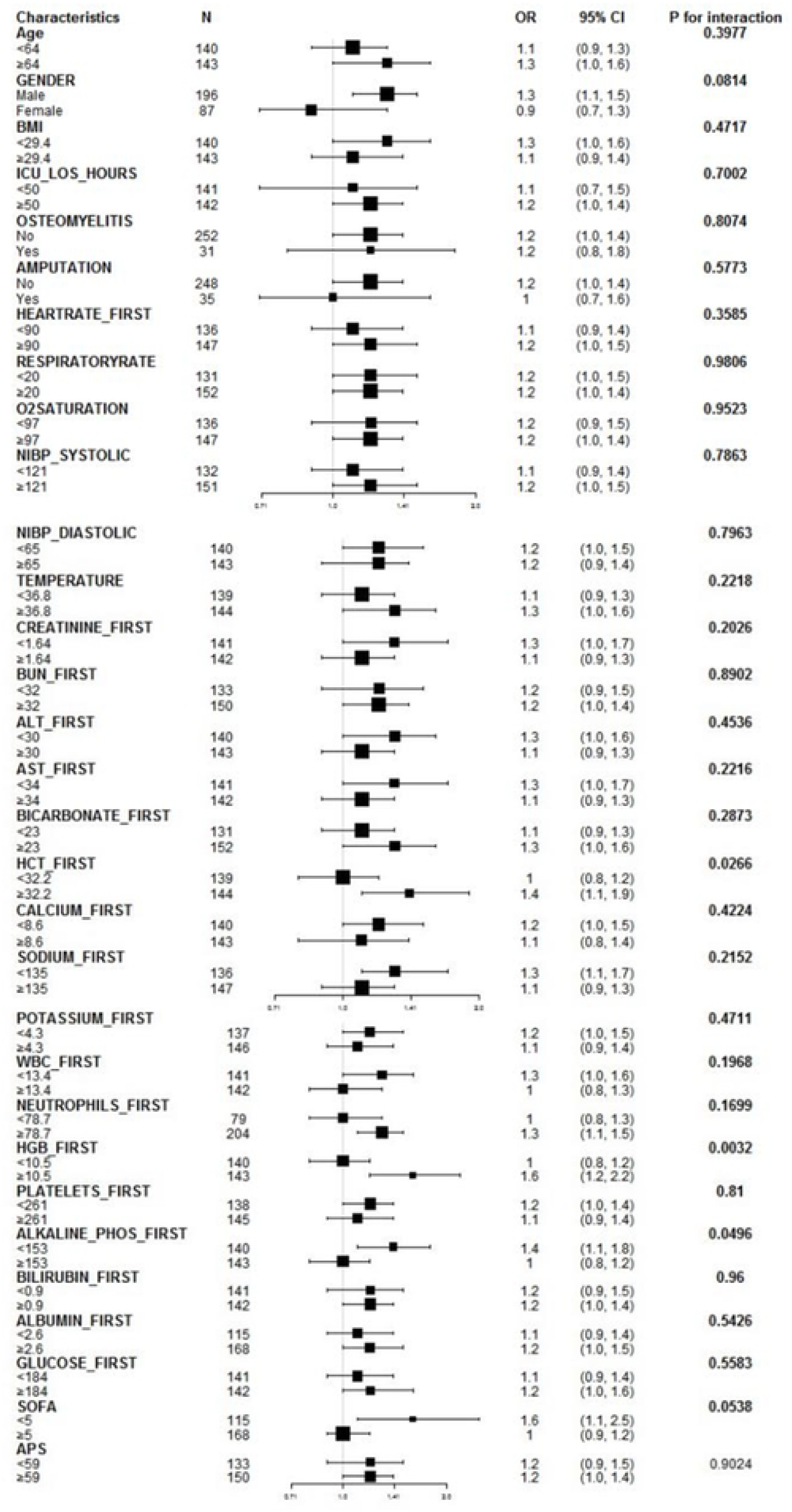
Subgroup analysis of in-hospital death and RDW.

## Discussion

Wound healing process of diabetes patients involved in many factors, which is slower than normal patients. Treatment of diabetes foot ulcer, especially ulcers with severe infections or osteomyelitis is still challenging. A multi-disciplinary team is required, including endocrinologist, plastic and reconstruction surgeon, nutritionist and interventional therapy doctors. Even though, a large number of patients face amputation or even death. Therefore, early prediction of patient prognosis is of great significance. This study confirms that RDW is an independent risk factor for diabetic foot mortality.

RDW is positively correlated with the mortality of diabetic foot patients. High RDW value means high mortality risk of diabetic foot, indicating that RDW can predict mortality and associated risk in DF patients. t is estimated that the survival rate is only 60% after 5 years^36^. Age, smoking, lower BMI, cardiovascular disease, kidney disease, osteomyelitis, femoral amputation, previous history of ulcer, patients with severe lesions, peripheral neuropathy, anemia and patients with HbA1c < 7% had been identified as independent predictors of mortality^36-41^. Hematological tests showed that neutrophil to lymphocyte ratio (NLR) and platelet to lymphocyte ratio (PLR) were reliable biomarkers for predicting mortality after DFU amputation^42, 43^. Meanwhile, male, smoking, amputation history, osteomyelitis history, peripheral artery disease, retinopathy, osteomyelitis, neuroischemic DFI, severe infection, leukocytosis, average ESR and average C-reactive protein (CRP) are also predictors of DF amputation^44-46^. Male gender, type 2 diabetes and smokers are significantly more other new diabetic foot ulcers or risk of relapse than those without these risk factors^47^.

Meanwhile, this study has several limitations. First, it is a single retrospective study and might be affected by selection bias. Second, in multivariate analysis, the absence of disease information may lead to bias. Finally, the follow-up time of these patients are different, and only hospital mortality is analyzed in this study, which might affect the mortality. Therefore, the explanation and mechanism of the relevance between RDW and mortality of DF need further study. Further research should be carried out to test this hypothesis.

Through the extraction and analysis of a large number of data, we found that RDW is a strong predictor of DF patients. The higher the RDW, the higher the risk of mortality in these patients. Further studies are needed to confirm the relationship between RDW and poor prognosis of DF.

## Data Availability

The clinical data used to support this study are available from Monitoring in Intensive Care Database III (MIMIC-III), which is publicly available and contains deidentified data from ICU patients. To obtain permission to access the database, researchers must complete the National Institutes of Health’s web-based course called Protecting Human Research Participants (certification number 29493483). The data used in our research adheres to the terms of this agreement, ensuring patient privacy and confidentiality. Researchers can obtain the MIMIC-III data set by following the specified access procedure on PhysioNet.

## Author contributions

Guangtao Huang and Tinggang Wang designed the study; Chunmei Gou and Guangtao Huang extracted the data; Chunmei Gou, Siyi Huang and Li Huang performed all the statistical analyses; Chunmei Gou drafed the paper. All authors revised and approved the final manuscript. Guangtao Huang and Tinggang Wang have contributed equally to this work.

## Supplementary Materials

The Supplementary file 1 used to extract data from MIMIC III.

